# Differing methodological quality but identical recommendations? - Assessment of methodological quality and content analysis of Clinical Practice Guidelines and Food-based Dietary Guidelines in early childhood allergy prevention

**DOI:** 10.1101/2023.05.26.23290529

**Authors:** Katharina Sieferle, Eva Maria Bitzer

**Author notes:** Corresponding author: Katharina Sieferle, Kunzenweg 21, 79117 Freiburg im Breisgau.

## Abstract

**Objective:** Recommendations on early childhood allergy prevention (ECAP) are found in clinical practice guidelines (CPG) and food-based dietary guidelines (FBDG). This study aims to compare the methodological quality and the content of recommendations in CPGs and FBDGs for ECAP.

**Study Design and Setting:** We assessed methodological quality of a sample of 36 guidelines (23 CPGs, 13 FBDGs), retrieved through extensive searching, with the Appraisal of Guidelines for Research and Evaluation tool (AGREE) II. On a subset of recommendations, we performed an in-depth analysis by type of intervention for direction of and strength of recommendation and level of evidence. Descriptive analysis was conducted with SPSS 27.

**Results:** CPGs scored higher than FBDGs in most AGREE domains (3, 4, 5 and 6). The 36 guidelines contain 280 recommendations on ECAP, with 68 addressing the introduction of complementary foods and allergenic foods. We found only slight differences between those recommendations in CPGs and FBDGs.

**Conclusion:** FBDGs on ECAP are of lower quality than CPGs. This does not affect their recommendations on the introduction of complementary foods and allergenic foods but may compromise their trustworthiness.

**What is new?:**

- Methodological quality of guidelines on ECAP is low, especially in FBDGs
- Recommendations on introduction of complementary feeding rarely vary
- Recommendations on introduction of potential allergenic foods show slight variation
- More attention is needed on the slight differences and the underlying evidence

## 1 Background

The prevalence of childhood allergies, including food allergies, eczema and asthma, is high, and associated with reduced quality of life and a high economic burden. The prevention of allergic diseases is therefore an important public health concern [1]. Recommendations on Early childhood allergy prevention (ECAP) are found in clinical practice guidelines (CPG) and food-based dietary guidelines (FBDG). The US Institute of Medicine defines CPGs as “statements that include recommendations intended to optimize patient care that are informed by a systematic review of evidence and an assessment of the benefits and harms of alternative care options” [2]. Especially in fields with rapidly evolving evidence like ECAP, CPGs and FBDGs are important to provide practitioners in ECAP and child nutrition (CN) with reliable guidance.

FBDGs and CPGs have the potential to improve healthcare quality and safety by translating research into practice [2,3]. However, considerable concern has been expressed by physicians, consumer groups, and other stakeholders about the quality of the processes supporting the development of FBDGs and CPGs [4]. Among the factors undermining the quality and trustworthiness are limitations in systematic reviews upon which CPGs are based, lack of transparency of development groups’ methodologies (particularly regarding evidence quality and strength of recommendation appraisals) and unmanaged conflicts of interest (COI) [5–7].

Even though guidelines should be based on systematic reviews, they are far from being “objective”, since making guideline recommendations always involves judgement, e.g. regarding the strengths and limitations of the evidence or the balance of benefits and harms [8]. The review of Perkin et al. shows for example, that different organizations and therefore different guidelines, interpret the evidence differently and come to diverging recommendation statements [9]. Moreover, CPGs and FBDGs come from different political, social, and economic fields, their developers have variable, but distinctive professional backgrounds, are exposed to different professional cultures, act under different economic premises and regulations. It seems reasonable to expect differences in the methodological approaches to develop such guidelines.

Diverging recommendations across guidelines might decrease the confidence in guidelines in general, if the reasoning leading to a recommendation statement is not transparent and no information regarding the developmental process is provided [2]. However, it has not been investigated systematically whether CPGs and FBDGs on ECAP comply with methodological standards in guideline development, and if not, whether the content of the recommendations is affected by different methodologies.

The objective of this study was to systematically assess the methodological quality of CPGs and FBDGs on ECAP and CN. We expect a clear understanding of the methodological quality of the guideline development, the strengths and weaknesses and the content of their recommendations.

## 2 Methods

This systematic synthesis of guidelines is reported according to the PRISMA (Preferred Reporting for Systematic Reviews and Meta-analyses) guidelines [10], when applicable, and a checklist is available on figshare (https://doi.org/10.6084/m9.figshare.22886672.v2).

### 2.1 Search strategy

We conducted a comprehensive search for national and international CPGs and FBDGs concerning ECAP and CN according to established recommendations for guideline retrieval [11]. Detailed information about the included databases, websites and the search strategy is available in the published study protocol [12]. A detailed list of (supra)national institutions included in the search can be found in tables A.1 and A.2 in appendix A.

### 2.2 Eligibility criteria

We considered all guidelines and recommendations about ECAP and CN, with infants or pregnant/breastfeeding women as target population, published since 2010 and valid at the time of search. Eligible were primary preventive interventions to decrease the onset of immediate or IgE mediated allergies, atopic eczema or asthma. We considered only CPGs and FBDGs, on a national or international level, and focusing on the topics asthma, atopic eczema, allergy prevention, food allergy and nutrition. Only publications in English or German were eligible.

Two study group members screened the retrieved records independently for their relevance according to the eligibility criteria and resolved any disagreements by discussion with the study group until consensus was reached.

### 2.3 Data extraction

The first author extracted basic data, which was cross-checked by another study group member. Among others, following data was extracted: Guideline title, first author, year of publication, country/scope, topic of the guideline, leading scientific societies, composition of guideline panel and document type (CPG, FBDG), and entered into a relational database.

### 2.4 Quality appraisal of guidelines

Two reviewers determined the quality of the included guidelines using the Appraisal of Guidelines for Research and Evaluation II (AGREE II) instrument [13]. Both assessors used the online training tools provided by the AGREE collaboration before conducting the assessment. AGREE II evaluates guideline methodology and quality and consists of 23 items covering six domains: (1) Scope and purpose, (2) Stakeholder involvement, (3) Rigour of development, (4) Clarity of presentation, (5) Applicability and (6) Editorial independence. Additionally, two overall assessment items are included: (7) A score from 1-7, indicating the general quality of the guideline and (8) the decision, whether the guideline can be recommended for use in practice. This “requires the user to make a judgement as to the quality of the guideline, taking into account the criteria considered in the assessment process” [13]. Each item of the AGREE II instrument is scored on a 7-point scale (1=strongly disagree to 7=strongly agree). We calculated a quality score for each domain, by summing up the scores of the individual items per domain and scaling the obtained score as a percentage of the maximum possible score for that domain:

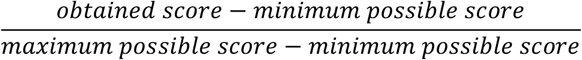

A higher domain score indicates a higher quality of the guideline in this domain. The six domain scores are independent and not aggregated into a single quality score [13].

We resolved discrepancies in scorings between the two reviewers by averaging the points if the scores differed by ≤1 point, and by discussion until consensus was reached if scores differed by two points or more.

### 2.5 Content analysis of guideline recommendations

We conducted a document analysis of relevant recommendation statements regarding ECAP. We derived codes for topics and time of intervention inductively from emerging themes in the guidelines, codes for the direction of recommendation, strength of recommendation (SoR) and level of evidence (LoE) were developed deductively based on the GRADE approach and adapted [14]. We grouped the topic of the interventions into 5 inductively derived groups: (1) nutrition-interventions in the child, (2) nutrition-interventions in the mother, (3) interventions regarding the environment, (4) medication and emollients and (5) other interventions. For the most frequent subgroup of recommendations, the introduction of complementary foods and potential allergens, we went deeper and compared the guidelines on the level of single recommendation statements regarding the direction and strength of the recommendation and the level of evidence.

Extraction of data and categorizing and coding of recommendations was conducted in MaxQDA.

### 2.6 Data storing and data management

We built a relational database containing data from the basic extraction, quality appraisal, and content analysis (MS Access ®). All databases and the software used are stored on secure servers of the University of Education Freiburg. All involved project members are bound by the Data Protection Act (DS-GVO) and subjected to confidentiality during the entire project phase and beyond.

### 2.7 Data analysis

We summarized general characteristics of the included guidelines using descriptive statistics. Quality scores for each AGREE II domain are presented using mean and standard deviation (SD). We conducted a t-test for independent samples to compare the mean domain scores for CPGs and FBDGs and a Mann-Whitney U-test for the variables that were not normally distributed. Statistical analysis of the extracted data was carried out with IBM SPSS Statistics software version 28.0.1.1 (14).

## 3 Results

### 3.1 Included guidelines

We identified a total of 2922 records and after removal of duplicates and title/abstract screening excluded 2676. After full-text screening of the remaining 246 records, 36 records were deemed eligible and included in our database (see flow-chart in figure 1). Of the 36 records, 23 are CPGs and 13 FBDGs. CPGs target a wider audience, with three CPGs published on international and two on European level, whereas included FBDGs are only published on national level (in n = 7 countries).

**Figure 1:**
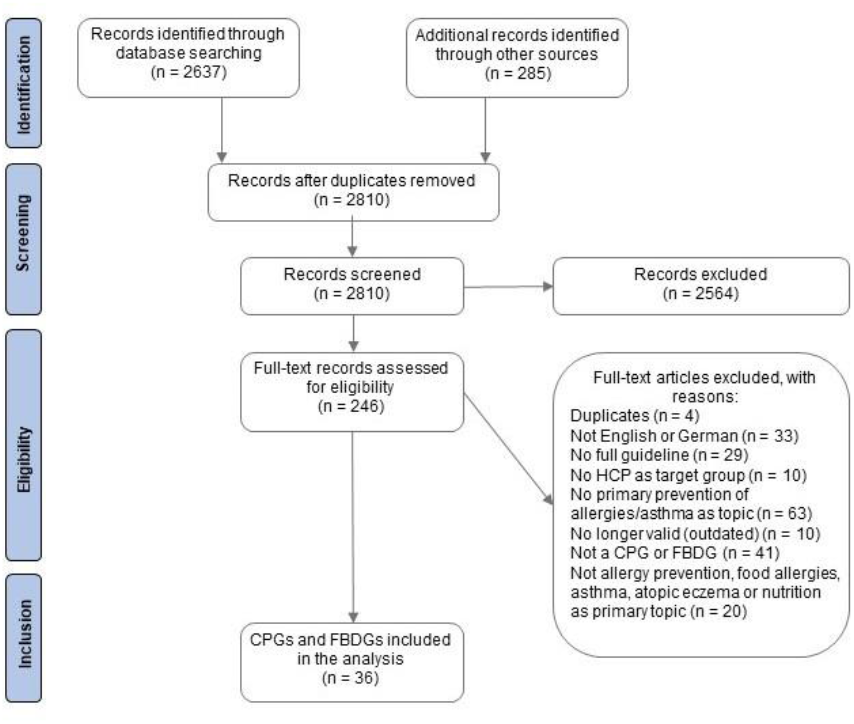
flow-chart of the records screened and full-texts retrieved

CPGs also cover a wider variety of topics, with nine guidelines addressing food allergies, six atopic eczema, five allergy prevention in general and three guidelines asthma. FBDGs are more homogeneous, with most FBDGs addressing child nutrition (n = 12) and only one directly addressing food allergies (for details see table A.3 in appendix A).

### 3.2 Guideline quality

Overall, the 36 guidelines score highest in the domains 4 “Clarity of Presentation” and 1 “Scope and Purpose” (79 % and 66 % respectively), and lowest in domains 5 “Applicability” and 6 “Editorial Independence” (24 % and 38 % respectively). The general quality of the guidelines (AGREE Item 7 “Overall Assessment”) is 63 %. Comparing CPGs and FBDGs, CPGs achieve almost consistently higher scores than FBDGs. We observe the largest difference in domain 6 “Editorial Independence” (Δ 33 points, 50 % vs. 17 %, p = 0.001), followed by domain 3 “Rigor of development” (Δ 17 points; 48 % vs. 31 %, p = 0.016). For details see figure 2, for results by guidelines table A.3 in appendix A.

**Figure 2:**
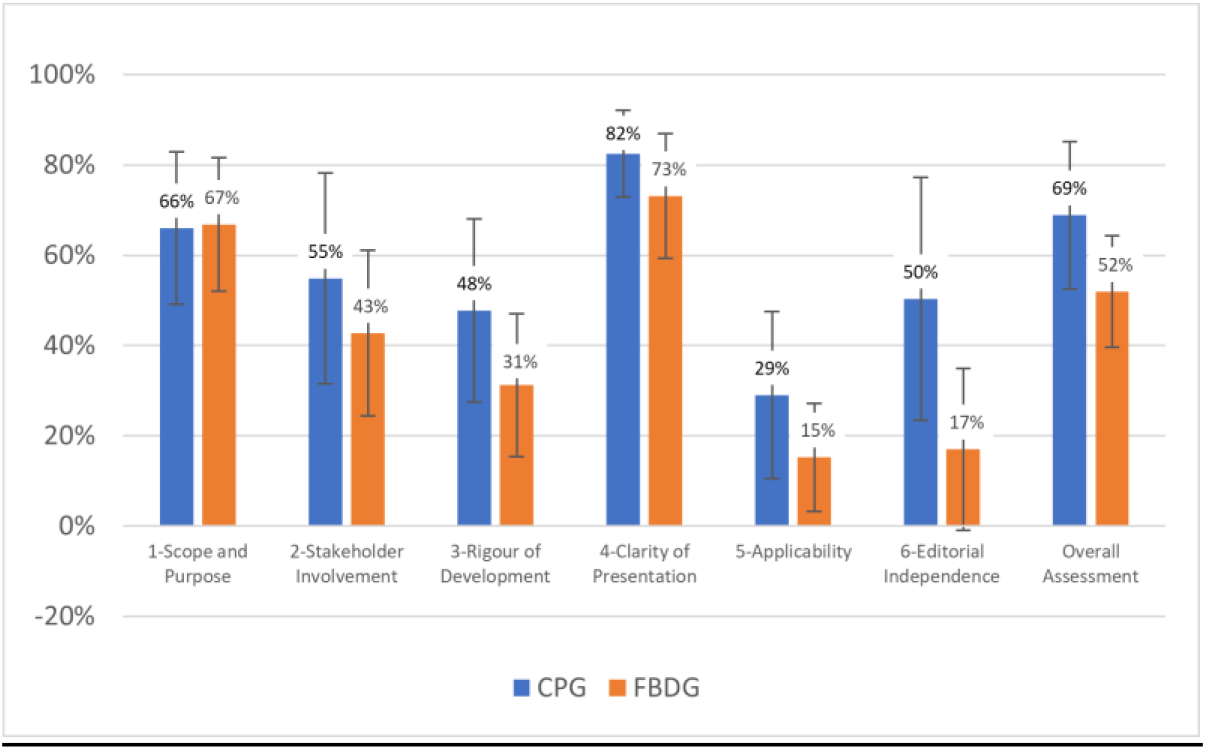
Comparison of standardised domain scores between CPGs and FBDGs (means and standard deviation)

### 3.3 Interventions addressed in guideline recommendation statements

The 36 guidelines contain 280 recommendations regarding ECAP. Most recommendations address nutrition-interventions for the child or mother (171 rs. 47). The most frequently addressed intervention is the introduction of complementary and allergenic foods (n = 68), followed by breastfeeding, maternal diet during pregnancy/lactation and breast milk substitutes and formula (n = 36, n = 33 and n = 30 respectively; for details see table 1).

**Table 1:**
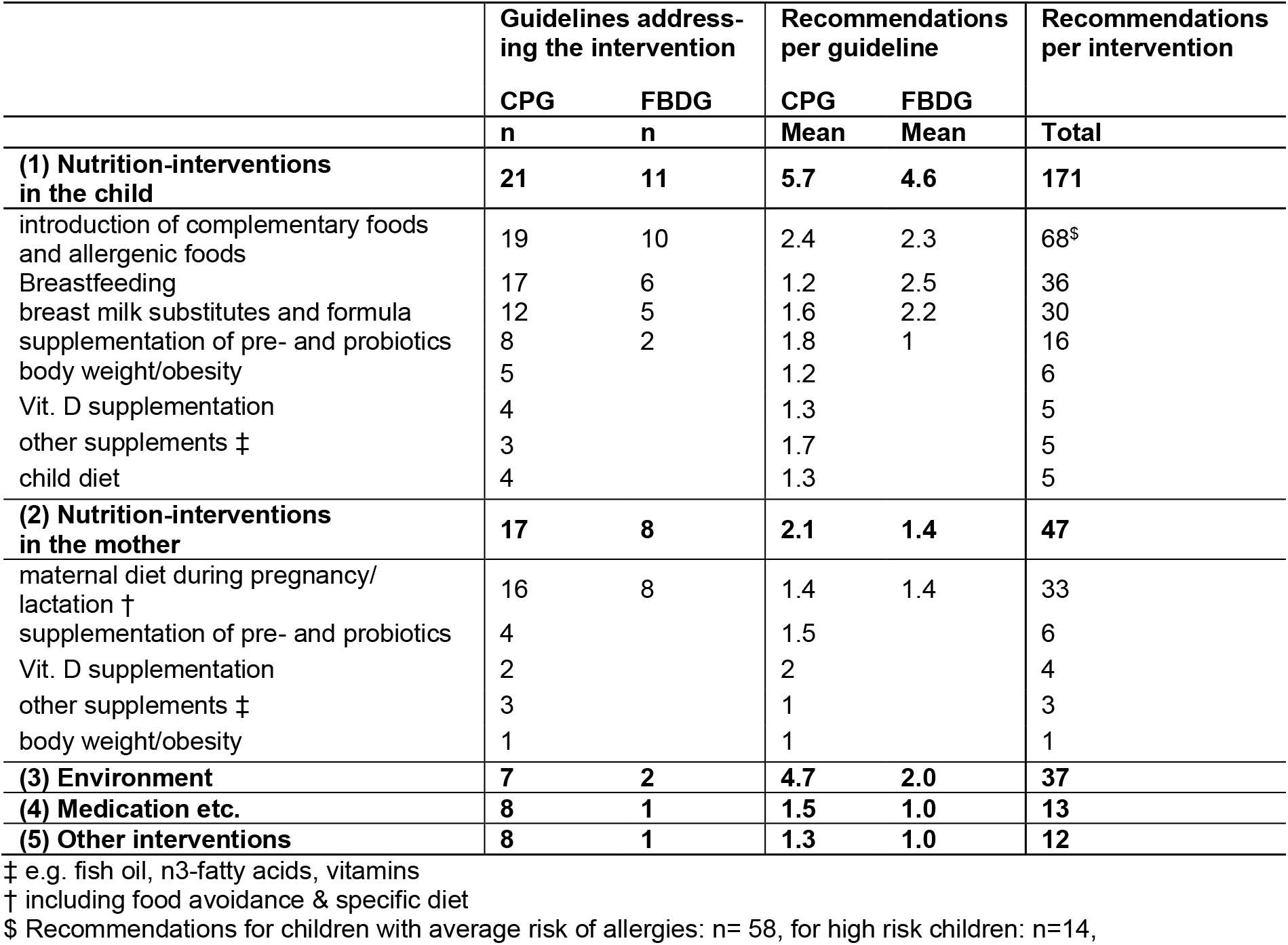
Interventions regarding allergy prevention in 23 CPGs and 13 FBDGs for children at average and high risk for allergies

|  | Guidelines addressing the intervention |  | Recommendations per guideline |  | Recommendations per intervention |
| --- | --- | --- | --- | --- | --- |
|  | CPG | FBDG | CPG | FBDG |  |
|  | n | n | Mean | Mean | Total |
| **(1) Nutrition-interventions in the child** | **21** | **11** | **5.7** | **4.6** | **171** |
| introduction of complementary foods and allergenic foods | 19 | 10 | 2.4 | 2.3 | 68<sup>\$</sup> |
| Breastfeeding | 17 | 6 | 1.2 | 2.5 | 36 |
| breast milk substitutes and formula | 12 | 5 | 1.6 | 2.2 | 30 |
| supplementation of pre- and probiotics | 8 | 2 | 1.8 | 1 | 16 |
| body weight/obesity | 5 |  | 1.2 |  | 6 |
| Vit. D supplementation | 4 |  | 1.3 |  | 5 |
| other supplements ‡ | 3 |  | 1.7 |  | 5 |
| child diet | 4 |  | 1.3 |  | 5 |
| **(2) Nutrition-interventions in the mother** | **17** | **8** | **2.1** | **1.4** | **47** |
| maternal diet during pregnancy/ lactation † | 16 | 8 | 1.4 | 1.4 | 33 |
| supplementation of pre- and probiotics | 4 |  | 1.5 |  | 6 |
| Vit. D supplementation | 2 |  | 2 |  | 4 |
| other supplements ‡ | 3 |  | 1 |  | 3 |
| body weight/obesity | 1 |  | 1 |  | 1 |
| **(3) Environment** | **7** | **2** | **4.7** | **2.0** | **37** |
| **(4) Medication etc.** | **8** | **1** | **1.5** | **1.0** | **13** |
| **(5) Other interventions** | **8** | **1** | **1.3** | **1.0** | **12** |

‡ e.g. fish oil, n3-fatty acids, vitamins
† including food avoidance & specific diet
\$ Recommendations for children with average risk of allergies: n= 58, for high risk children: n=14,

CPGs cover a wider range of ECAP interventions compared to FBDGs, and often provide more recommendations per intervention and guideline. For interventions in the child nearly all CPGs and FBDGs address the introduction of complementary foods and allergenic foods, half of the guidelines address breast milk substitutes and formula, whereas child body weight, Vitamin D and other supplements are covered in CPGs only. 17 of 23 CPGS (and 8 of 13 FBDGs) address interventions targeted at the mother, usually maternal diet during pregnancy/lactation (see table 1). A third of the CPGs, but only two FBDGs include recommendations on interventions regarding the environment and medication (pet ownership, motor-vehicle emissions, vaccination [15] and exposure to tobacco smoke [15,16]). The average number of recommendations per guideline on a given intervention varies between CPGs and FBDGs depending on the type of intervention. CPGs provide more recommendations on introduction of complementary food, maternal interventions, and environmental interventions, whereas FBDGs offered more recommendations on breastfeeding and breast milk substitution (see table 1).

### 3.4 Recommendations on the most commonly addressed intervention: introduction of complementary foods and potential allergenic foods

Do CPGs and FBDGs agree on their recommendations on interventions covered in both types of guidelines? Of 68 single recommendations on the introduction of complementary foods and potential allergenic foods (weaning) 58 are directed at children at average risk for allergies, 14 recommendations address high risk children (4 recommendations address both groups). We condensed the 58 recommendations on weaning in average risk children to 26 content identical recommendations (see table 2). (See Table A.4 in appendix A for recommendations for high-risk children).

**Table 2:**
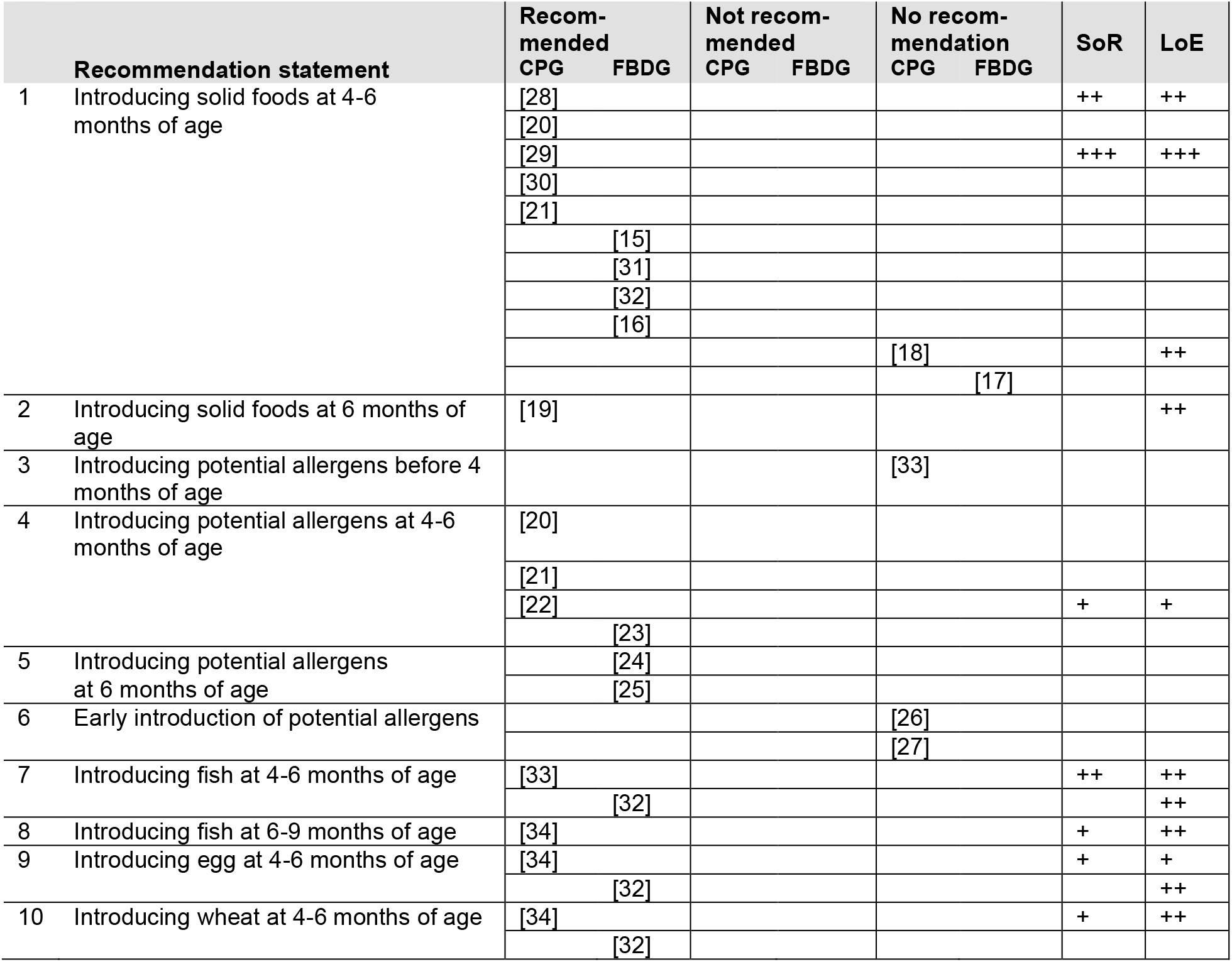
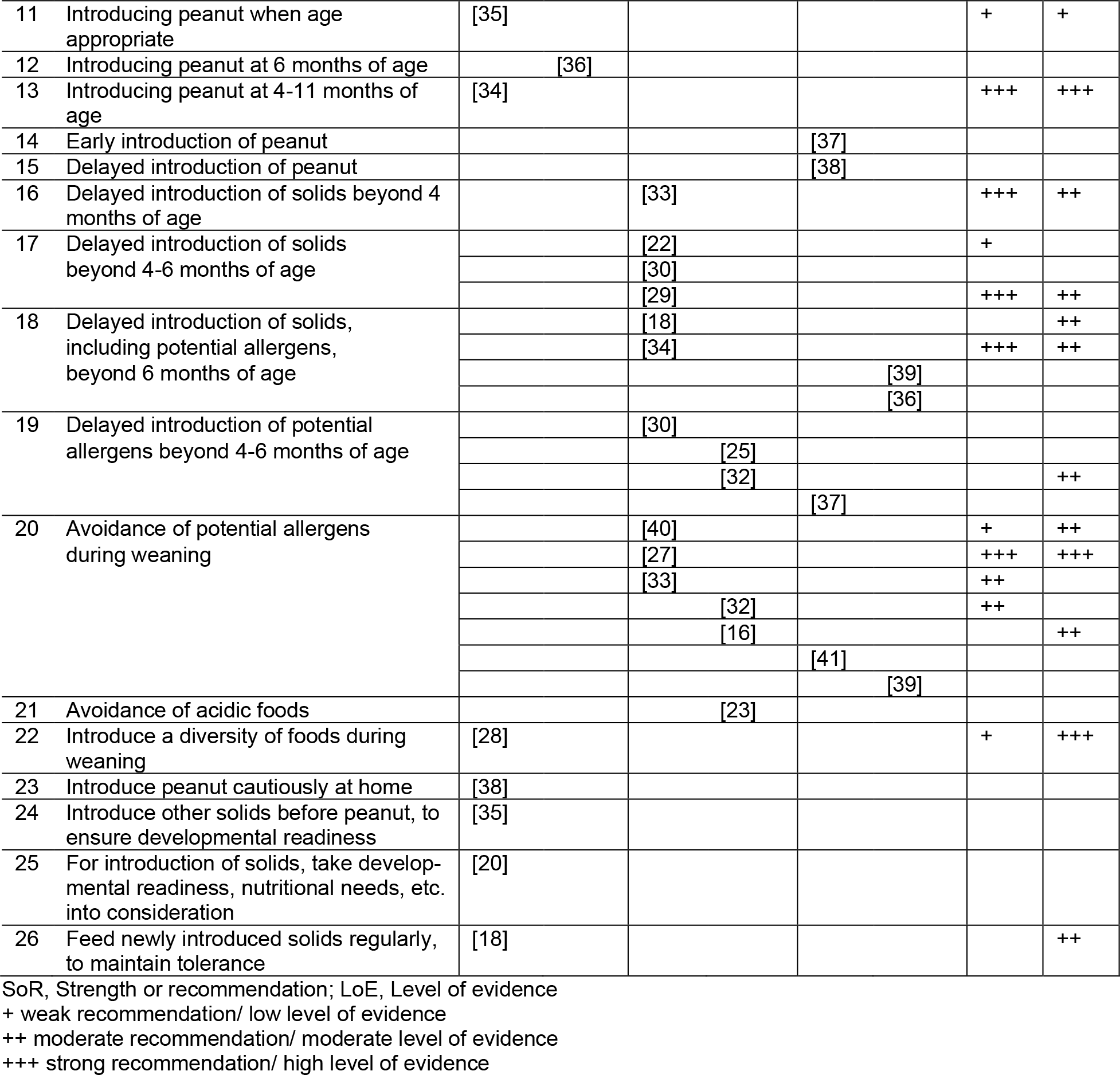
26 content identical recommendation statements regarding the introduction of complementary foods and allergenic foods in CPGs and FBDGs for average risk children (direction and strength of recommendation and level of evidence)

Information on the LoE is available in 23 of 58 recommendations (40%), 18 are accompanied by a grading of the SoR (31%), 15 of 58 recommendations (26%) are presented with LoE and SoR.

Most guidelines agree on the introduction of complementary foods: they recommend introducing solids between four and six months of age, and/or advice against delaying the introduction beyond that age. Two guidelines, published in 2015 and 2013 [17,18] make no recommendation on the appropriate timing yet, because “inducing tolerance by introducing solid foods at four to six months of age is currently under investigation and cannot be recommended at this time”, and one guideline, published 2011, recommends introducing solids only at six months of age “Current UK guidelines based on recommendations from the WHO recommend that weaning should start at six months.” [19].

There is slightly less agreement regarding the introduction of potentially allergenic foods in general: among the 20 recommendations some focus on specific timepoints of introduction, some advise for early, or against a delayed introduction. Guidelines vary in recommending the introduction between four to six months of age [20–23], after six month [24,25], and two guidelines refrain explicitly from a recommendation but do not provide formal assessment of LoE or SoR [26,27]. The few recommendations that address the introduction of single allergens, e.g. peanut, egg, wheat and fish, do have nuanced differences.

By visual inspection of the table we could not find a convincing association between the content of the recommendations and the use of SoR or LoE ratings.

The much shorter table on recommendations directed at high-risk children depicts a similar picture: more or less nuanced discrepancies between CPGs and FBDGs and no apparent association between SoR or LoE ratings and the content of the recommendation (see table A.4 in appendix A).

## 4 Discussion

The aim of this study is to systematically assess the methodological quality of CPGs and FBDGs on ECAP and CN. Our expectation is to obtain a comprehensive understanding of the methodological rigour employed in the guideline development, to identify their strengths and weaknesses, and to explore the content of their recommendation.

Our findings indicate that the quality of CPGs and FBDGs on ECAP and CN differs significantly, with especially low scores in AGREE-domains 3 “Rigour of Development”, 5 “Applicability” and 6 “Editorial Independence”. These domains focus on the methodological quality of the guideline development, including evidence search and synthesis, COI management and guideline implementation. Other studies have shown similar deficits in the methodological quality of guideline development [42,43], but not in direct comparison of guidelines from two different subject fields.

CPGs generally scored higher than FBDGs, possibly because methodological standards for guideline development differ for CPGs and FBDGs and are not as widely used in FBDGs yet [44]. Differences were especially prevalent in domains 3 and 6 and could be seen as an urgent need to implement higher methodological standards in the development of FBDGs, even when acknowledging that the methods used to develop and assess CPGs are not always suitable for FBDGs, and the questions FBDGs want to answer [45].

In our sample, the guidelines with high methodological quality according to AGREE II were developed by organisations such as the AWMF, SIGN, GINA, NIAID, AAD and EAACI [19,22,27,33,46–48]. These medical organisations are often very active in the scientific development and testing of methods to produce high quality guidelines, have published detailed guidance on guideline development, or use the tools GRADE [14] and AGREE II, therefore ensuring a higher methodological standard.

Although the methodological quality varied considerably across the guidelines, the recommendation statements exhibited only minor, in some cases rather nuanced discrepancies. Most guidelines recommended introducing complementary foods, including allergenic foods, between four and six months of age, and advised against delaying the introduction of any food items. However, a few guidelines made other recommendations, e.g., the SIGN guideline on the management of atopic eczema from 2011, which indirectly recommends the introduction of complementary foods at 6 months [19]. The observed minor differences in recommendations may partly be explained by the year of publication and the evidence used and available at that time.

Only a quarter of the recommendation statements were given a formal grading of the SoR and LoE, usually attributed to higher methodological rigour and quality. However, this turned out not to be relevant when it came to the question “What to recommend?”, since we saw no convincing associations between those gradings and the content of the recommendations.

How did FBDGs reach recommendations quite similar to CPGs despite their significantly lower methodological quality? Is the higher methodological standard even necessary to provide clear recommendations on ECAP? We caution against such a conclusion for several reasons: 1) It might be the case in ECAP, that the paradigm shift from avoidance of allergens to early and sustained exposure has gained widespread acceptance among clinicians and nutritionists. Then, providing guideline recommendations on ECAP would simply reflect common knowledge, without the need to prove the evidence again (like buckling up in cars). But how would we (or a guideline panel) asses the level of “unity”, “common knowledge” or “controversy” to decide for or against a more elaborate methodological process? 2) Our in-depth assessment of the recommendation statements focused solely on the topic of introduction of complementary foods and potential allergenic foods. For other ECAP interventions, such as formula and breast milk substitutes or pre- and probiotic supplementation, the situation could be much different. Lower methodological quality, especially lack of information on COI management, as reflected in AGREE-domain 6, may play a substantial role here [49,50]. 3) Our study indicates that it takes a lot of resources, efforts, and manpower to check if guidelines developed with lower quality recommend roughly the same as higher quality guidelines. So even if lower quality guideline recommendations do not differ substantially from high quality guidelines, low methodological quality and little transparency can lead to distrust in the recommendations and hinder the application of the guideline in clinical / extraclinical practice [2].

We would like to point at another issue: AGREE II does assess the rigor of development, which includes the search for evidence and formulating of the recommendations, but does not take the evidence base of the recommendations in detail into account. A close look on the type of sources used to justify the recommendation statements on ECAP would be interesting and worthwhile. An exploration if guidelines of varying rigor of development and management of conflicts of interest use similar evidence (i.e. systematic reviews, high quality primary studies) would be a logical next step.

In conclusion, this investigation highlights deficits in guideline development, especially in FBDGs. Our results contribute to a better understanding of the importance of the methodological rigor and management of evidence and COI in guidelines. Although the recommendations concerning the introduction of complementary feeding in our sample showed low variability, since childhood allergies are an important public health concern, and effective allergy prevention in early childhood available, feasible but not fully implemented on population level, a higher methodological quality of guidelines concerning ECAP should be promoted to increase confidence in the recommendations.

## Data Availability

All data produced in the present study are contained in the manuscript or available upon reasonable request to the corresponding author.

## Acknowledgements

We thank Nina Wünst for her contribution to the data collection and extraction.

## Author Contributions

EMB: Conceptualization, Funding acquisition, Methodology, Project administration, Supervision, Writing – review & editing; KS: Conceptualization, Data curation, Formal analysis, Methodology, Visualisation, Writing – original draft, Writing – review & editing

## References

[1] Pawankar R. Allergic diseases and asthma: a global public health concern and a call to action. World Allergy Organ J 2014;7(1):12. https://doi.org/10.1186/1939-4551-7-12.

[2] Graham R. Clinical practice guidelines we can trust. Washington, DC: National Academies Press; 2011.

[3] Qaseem A, Forland F, Macbeth F, Ollenschläger G, Phillips S, van der Wees P. Guidelines International Network: Toward international standards for clinical practice guidelines. Ann Intern Med 2012;156(7):525–31. https://doi.org/10.7326/0003-4819-156-7-201204030-00009.

[4] Bindslev JBB, Schroll JB, Gøtzsche PC, Lundh A. Underreporting of conflicts of interest in clinical practice guidelines: Cross sectional study. BMC Med Ethics 2013;14:19. https://doi.org/10.1186/1472-6939-14-19.

[5] Hansen C, Lundh A, Rasmussen K, Gøtzsche PC, Hróbjartsson A. Financial conflicts of interest and outcomes and quality of systematic reviews. Cochrane Database Syst Rev 2017;365(9465):1159. https://doi.org/10.1002/14651858.MR000047.

[6] Lundh A, Lexchin J, Mintzes B, Schroll JB, Bero LA. Industry sponsorship and research outcome. Cochrane Database Syst Rev 2017;2:MR000033. https://doi.org/10.1002/14651858.MR000033.pub3.

[7] Shekelle PG. Clinical practice guidelines: What’s next? JAMA 2018;320(8):757–8. https://doi.org/10.1001/jama.2018.9660.

[8] Arbeitsgemeinschaft der Wissenschaftlichen Medizinischen Fachgesellschaften (AWMF) - Ständige Kommission Leitlinien. AWMF-Regelwerk “Leitlinien”: 1. Auflage 2012 2012.

[9] Perkin MR, Togias A, Koplin J, Sicherer S. Food Allergy Prevention: More Than Peanut. The Journal of Allergy and Clinical Immunology: In Practice 2020;8(1):1–13. https://doi.org/10.1016/j.jaip.2019.11.002.

[10] Page MJ, McKenzie JE, Bossuyt PM, Boutron I, Hoffmann TC, Mulrow CD et al. The PRISMA 2020 statement: An updated guideline for reporting systematic reviews. PLoS Med 2021;18(3):e1003583. https://doi.org/10.1371/journal.pmed.1003583.

[11] Blümle A, Sow D, Nothacker M, Schaefer C, Motschall E, Boeker M et al. Manual systematische Recherche für Evidenzsynthesen und Leitlinien. Albert-Ludwigs-Universität Freiburg; 2019.

[12] Sieferle K, Schaefer C, Bitzer EM. Management of evidence and conflict of interest in guidelines on early childhood allergy prevention and child nutrition: study protocol of a systematic synthesis of guidelines and explorative network analysis [version 1; peer review: awaiting peer review]. F1000Res 2022;11(1290). https://doi.org/10.12688/f1000re-search.123571.1.

[13] Brouwers M, Kho ME, Browman GP, Burgers JS, Cluzeau F, Feder G, Fervers B, Graham ID, Grimshaw J, Hanna S, Littlejohns P, Makarski J, Zitzelsberger L for the AGREE Next Steps Consortium. AGREE II: Advancing guideline development, reporting and evaluation in healthcare. Can Med Assoc J. 2010. https://doi.org/10.1503/cmaj.090449.

[14] Schünemann H, Brożek J, Guyatt G, Oxman A. GRADE Handbook: for grading the quality of evidence and the strength of recommendations using the GRADE approach. [April 06, 2020]; Available from: https://gdt.gradepro.org/app/handbook/handbook.html.

[15] Koletzko B, Bauer C-P, Cierpka M, Cremer M, Flothkötter M, Graf C et al. Ernährung und Bewegung von Säuglingen und stillenden Frauen: Aktualisierte Handlungsempfehlungen von “Gesund ins Leben - Netzwerk Junge Familie”, eine Initiative von IN FORM. Monatsschr Kinderheilkd 2016;164(S5):433–57.

[16] Schweizerische Gesellschaft für Ernährung. Ernährung des Säuglings im ersten Lebensjahr. [December 12, 2022]; Available from: http://www.sge-ssn.ch/media/Merk-blatt_Ernaehrung_des_Saeuglings_im_ersten_Lebensjahr-2019.pdf.

[17] Alberta Health Services. Nutrition Guideline Healthy Infants and Young Children Introduction of Complementary Foods: Applicable to: Nurses, Physicians and Other Health Professionals. [December 12, 2022].

[18] Chan ES, Cummings C. Dietary exposures and allergy prevention in high-risk infants: A joint statement with the Canadian Society of Allergy and Clinical Immunology. Paediatr Child Health 2013;18(10):545–54. https://doi.org/10.1093/pch/18.10.545.

[19] Scottish Intercollegiate Guidelines Network. Management of atopic eczema in primary care: A national clinical guideline. (SIGN Guideline No 125). Edinburgh; 2011.

[20] Chan AW, Chan JK, Tam AY, Leung TF, Lee TH. Guidelines for allergy prevention in Hong Kong. Hong Kong Med J 2016;22(3):279–85. https://doi.org/10.12809/hkmj154763.

[21] Di Mauro G, Bernardini R, Barberi S, Capuano A, Correra A, De’ Angelis GL et al. Prevention of food and airway allergy: consensus of the Italian Society of Preventive and Social Paediatrics, the Italian Society of Paediatric Allergy and Immunology, and Italian Society of Pediatrics. World Allergy Organ J 2016;9:1–28. https://doi.org/10.1186/s40413-016-0111-6.

[22] Boyce JA, Assa’ad AH, Burks W, Jones SM, Sampson HA, Wood RA et al. Guidelines for the diagnosis and management of food allergy in the United States: report of the NIAID-sponsored expert panel. J Allergy Clin Immunol 2010;126(6 Suppl):S1–58. https://doi.org/10.1016/j.jaci.2010.10.007.

[23] Pérez-Escamilla R, Segura-Pérez S, Lott M. Feeding Guidelines for Infants and Young Toddlers: A Responsive Parenting Approach. Nutrition Today 2017;52(5):223–231. https://doi.org/10.1097/NT.0000000000000234.

[24] Health Canada, Canadian Paediatric Society, Dietitians of Canada, Breastfeeding Committee for Canada. Nutrition for Healthy Term Infants: Recommendations from Six to 24 Months. [December 12, 2022]; Available from: https://www.canada.ca/en/health-can-ada/services/canada-food-guide/resources/infant-feeding/nutrition-healthy-term-infants-recommendations-birth-six-months/6-24-months.html.

[25] Health Canada, Canadian Paediatric Society, Dietitians of Canada, Breastfeeding Committee for Canada. Nutrition for healthy term infants: Recommendations from birth to six months. [December 12, 2022]; Available from: https://www.canada.ca/en/health-can-ada/services/canada-food-guide/resources/infant-feeding/nutrition-healthy-term-infants-recommendations-birth-six-months.html#a4.

[26] Clark AT, Skypala I, Leech SC, Ewan PW, Dugué P, Brathwaite N et al. British Society for Allergy and Clinical Immunology guidelines for the management of egg allergy. Clin Exp Allergy 2010;40(8):1116–29. https://doi.org/10.1111/j.1365-2222.2010.03557.x.

[27] Scottish Intercollegiate Guidelines Network, British Thoracic Society. SIGN158 British Guideline on the management of asthma: A national clinical guideline. (SIGN Guideline No 158). Edinburgh; 2019.

[28] Wollenberg A, Barbarot S, Bieber T, Christen-Zaech S, Deleuran M, Fink-Wagner A et al. Consensus-based European guidelines for treatment of atopic eczema (atopic dermatitis) in adults and children: part I. J Eur Acad Dermatol Venereol 2018;32(5):657–82. https://doi.org/10.1111/jdv.14891.

[29] Lee BW, Aw MM, Chiang WC, Daniel M, George GM, Goh EN et al. Academy of medicine, Singapore-Ministry of Health clinical practice guidelines: management of food allergy. Singapore Med J 2010;51(7):599–607.

[30] Ebisawa M, Ito K, Fujisawa T. Japanese guidelines for food allergy 2017. Allergol Int 2017;66(2):248–64. https://doi.org/10.1016/j.alit.2017.02.001.

[31] National Institute for Health and Welfare in Finland. EATING TOGETHER - food recommendations for families with children. [December 09, 2022]; Available from: http://www.julkari.fi/bitstream/handle/10024/137770/URN_ISBN_978-952-343-264-2.pdf?sequence=1&isAllowed=y.

[32] Österreichische Gesellschaft für Kinder-und Jugendheilkunde. Österreichische Beikostempfehlungen: Richtig essen von Anfang an! [December 23, 2022]; Available from: https://www.richtigessenvonanfangan.at/down-load/0/0/ebb0d4ccc1fb0d8300146bc8a90b3c88fd4165fb/fileadmin/Redakteure_RE-VAN/user_upload/Beikostempfehlungen_Expertenversion_24-04-2013.pdf.

[33] Schäfer T, Bauer C-P, Beyer K, Bufe A, Friedrichs* F, Gieler U et al. S3-Leitlinie Aller-gieprävention - Update 2014. [March 10, 2020]; Available from: https://www.awmf.org/uploads/tx_szleitlinien/061-016l_S3_Allergiepr%C3%A4vention_2014-07-abgelaufen.pdf.

[34] Recto MST, Genuino MLG, Castor MAR, Casis-Hao RJ, Tamondong-Lachica DR, Sales MIV et al. Dietary primary prevention of allergic diseases in children: the Philippine guidelines. Asia Pac Allergy 2017;7(2):102–14. https://doi.org/10.5415/apal-lergy.2017.7.2.102.

[35] Togias A, Cooper SF, Acebal ML, Assa’ad AH, Baker JR, Beck LA et al. Addendum guidelines for the prevention of peanut allergy in the United States: Report of the National Institute of Allergy and Infectious Diseases-sponsored expert panel. J Allergy Clin Immunol 2017;139(1):29–44. https://doi.org/10.1016/j.jaci.2016.10.010.

[36] National Health and Medical Research Council. Australian Dietary Guidelines: Providing the scientific evidence for healthier Australian diets. [December 12, 2022]; Available from: http://www.nhmrc.gov.au/guidelines-publications/n55.

[37] Greer FR, Sicherer SH, Burks AW. The Effects of Early Nutritional Interventions on the Development of Atopic Disease in Infants and Children: The Role of Maternal Dietary Restriction, Breastfeeding, Hydrolyzed Formulas, and Timing of Introduction of Allergenic Complementary Foods. Pediatrics 2019. https://doi.org/10.1542/peds.2019-0281.

[38] Stiefel G, Anagnostou K, Boyle RJ, Brathwaite N, Ewan PW, Fox AT et al. BSACI guideline for the diagnosis and management of peanut and tree nut allergy. Clin Exp Allergy 2017;47(6):719–39. https://doi.org/10.1111/cea.12957.

[39] National Health and Medical Research Council. Infant Feeding Guidelines: Information for health workers. [December 09, 2022].

[40] Sampson HA, Aceves S, Bock SA, James J, Jones SM, Lang D et al. Food allergy: a practice parameter update-2014. J Allergy Clin Immunol 2014;134(5):1016–25.e43. https://doi.org/10.1016/j.jaci.2014.05.013.

[41] Lansang P, Lam JM, Marcoux D, Prajapati VH, Spring S, Lara-Corrales I. Approach to the Assessment and Management of Pediatric Patients With Atopic Dermatitis: A Consensus Document. Section III: Treatment Options for Pediatric Atopic Dermatitis. J Cutan Med Surg 2019;23(5_suppl):19S–31S. https://doi.org/10.1177/1203475419882647.

[42] Bhatt M, Nahari A, Wang P-W, Kearsley E, Falzone N, Chen S et al. The quality of clinical practice guidelines for management of pediatric type 2 diabetes mellitus: a systematic review using the AGREE II instrument. Syst Rev 2018;7(1):193. https://doi.org/10.1186/s13643-018-0843-1.

[43] Chiappini E, Bortone B, Galli L, Martino M de. Guidelines for the symptomatic management of fever in children: systematic review of the literature and quality appraisal with AGREE II. BMJ Open 2017;7(7):e015404. https://doi.org/10.1136/bmjopen-2016-015404.

[44] EFSA Panel on Dietetic Products, Nutrition and Allergies. Scientific Opinion on establishing Food-Based Dietary Guidelines. EFS2 2010;8(3). https://doi.org/10.2903/j.efsa.2010.1460.

[45] Bero LA, Norris SL, Lawrence MA. Making nutrition guidelines fit for purpose. BMJ 2019;365:1579. https://doi.org/10.1136/bmj.l1579.

[46] Global Initiative for Asthma. Global Strategy for Asthma Management and Prevention: Updated 2020 2020.

[47] Eichenfield LF, Tom WL, Chamlin SL, Feldman SR, Hanifin JM, Simpson EL et al. Guidelines of care for the management of atopic dermatitis: Section 1. Diagnosis and assessment of atopic dermatitis. Journal of the American Academy of Dermatology 2013;70(2):338–51. https://doi.org/10.1016/j.jaad.2013.10.010.

[48] Muraro A, Halken S, Arshad SH, Beyer K, Dubois AEJ, Du Toit G et al. EAACI food allergy and anaphylaxis guidelines. Primary prevention of food allergy. Allergy 2014;69(5):590–601. https://doi.org/10.1111/all.12398.

[49] Worl Health Organization. Marketing of breast-milk substitutes: national implementaion of the international code. status report 2018 2018.

[50] Rollins N, Piwoz E, Baker P, Kingston G, Mabaso KM, McCoy D et al. Marketing of commercial milk formula: a system to capture parents, communities, science, and policy. The Lancet 2023;401(10375):486–502. https://doi.org/10.1016/S0140-6736(22)01931-6.

[51] Pedersen S, Hurd SS, Lemanske RF JR, Becker AB, Zar HJ, Sly PD et al. Global strategy for the diagnosis and management of asthma in children 5 years and younger. Pediatr Pulmonol 2011;46(1):1–17. https://doi.org/10.1002/ppul.21321.

[52] Rajagopalan M, De A, Godse K, Krupa Shankar DS, Zawar V, Sharma N et al. Guidelines on Management of Atopic Dermatitis in India: An Evidence-Based Review and an Expert Consensus. Indian J Dermatol 2019;64(3):166–81. https://doi.org/10.4103/ijd.IJD_683_18.

[53] Nowak-Węgrzyn A, Chehade M, Groetch ME, Spergel JM, Wood RA, Allen KJ et al. International consensus guidelines for the diagnosis and management of food protein-induced enterocolitis syndrome: Executive summary-Workgroup Report of the Adverse Reactions to Foods Committee, American Academy of Allergy, Asthma & Immunology. J Allergy Clin Immunol 2017;139(4):1111–1126.e4. https://doi.org/10.1016/j.jaci.2016.12.966.

[54] Nordic Council of Ministers. Nordic Nutrition Recommendations 2012: Integrating nutrition and physical activity. 5th ed. Copenhagen: Nordic Council of Ministers; 2014.

[55] Koletzko B, Cremer M, Flothkötter M, Graf C, Hauner H, Hellmers C et al. Ernährung und Lebensstil vor und während der Schwangerschaft - Handlungsempfehlungen des bundesweiten Netzwerks Gesund ins Leben. Geburtshilfe Frauenheilkd 2018;78(12):1262–82. https://doi.org/10.1055/a-0713-1058.

[56] Bürklin S, Relats C, Herzog R, Stalder K, Roduit C, Fischer I et al. Ernährungsberatung bei Kindern mit IgE-vermittelten Nahrungsmittelallergien: Eine Praxisleitlinie; 2019.

