## Appendix A for "Differing methodological quality but identical recommendations? - Assessment of methodological quality and content analysis of Clinical Practice Guidelines and Food-based Dietary Guidelines in early childhood allergy prevention"

Table A. 1 Food and Agriculture Organization of the United Nations (FAO) directory for institutions reporting on FBDGs

| Europe | Institutional contacts |
| --- | --- |
| Albania | [Ministry of Health](http://www.shendetesia.gov.al/) |
| Austria | [Ministry of Health](http://www.bmg.gv.at/) |
| Belgium | [Federal Public Service Health, Food Chain Safety and](http://www.health.belgium.be/eportal/index.htm) [Environment](http://www.health.belgium.be/eportal/index.htm) |
| Bosnia and Herzegovina | [Institute of Public Health of Federation of Bosnia and](http://www.zzjzfbih.ba/)  [Herzegovina](http://www.zzjzfbih.ba/) |
| Bulgaria | [Ministry of Health](http://www.mh.government.bg/bg/) |
| Croatia | [Ministry of Health](http://www.zdravlje.hr/) |
| Cyprus | [Ministry of Health](http://www.moh.gov.cy/moh/moh.nsf/index_gr/index_gr?OpenDocument) |
| Denmark | [Ministry of Food, Agriculture and Fisheries](http://www.altomkost.dk/Services/Kontakt/forside.htm) |
| Estonia | [National Institute for Health Development](http://www.toitumine.ee/trukised/?type=13997) |
| Finland | [National Nutrition Council](http://www.ravitsemusneuvottelukunta.fi/portal/en/nutrition%2Brecommendations/) |
| France | [Ministry of Health](http://social-sante.gouv.fr/) |
| Georgia | [National Centre for Disease Control and Public Health](http://www.ncdc.ge/?lang=eng) |
| Germany | [German Nutrition Society](http://www.dge.de/modules.php?name=Content&pa=showpage&pid=12) |
| Greece | [National and Kapodistrian University of Athens, School of](http://www.nut.uoa.gr./contactENG.html) [Medicine - WHO Collaborating Center for Food and Nutrition](http://www.nut.uoa.gr./contactENG.html)  [Policies](http://www.nut.uoa.gr./contactENG.html) |
| Hungary | [National Institute for Food and Nutrition Science](http://www.oeti.hu/index.php?m1id=20&m2id=207) |
| Iceland | [The Directorate of Health](http://www.landlaeknir.is/english/) |
| Ireland | [Department of Health](https://www.healthpromotion.ie/) |
| Israel | [Ministry of Health](http://www.health.gov.il/Subjects/FoodAndNutrition/Nutrition/Adequate_nutrition/Pages/default.aspx) |
| Italy | [Research Centre on Food and Nutrition](http://sito.entecra.it/portale/index2.php?lingua=IT&access_flag=0) |
| Latvia | [Ministry of Health](http://www.vm.gov.lv/) |
| Malta | [The Health Promotion and Disease Prevention Directorate,](https://ehealth.gov.mt/HealthPortal/default.aspx) [Parliamentary Secretariat for Health](https://ehealth.gov.mt/HealthPortal/default.aspx) |
| Netherlands | [Netherlands Nutrition Centre](http://www.voedingscentrum.nl/) |
| Norway | [Directorate of Health](http://www.helsedirektoratet.no/Sider/default.aspx) |
| Poland | [National Food and Nutrition Institute](http://www.izz.waw.pl/en/) |
| Portugal | [Faculty of Food Sciences and Nutrition, Porto University](http://www.fcna.up.pt/) |
| Romania | [National Food and Nutrition Committee, Ministry of Health](http://www.ms.gov.ro/?pag=1) |
| Slovenia | [National Institute of Public Health](http://www.nijz.si/) |
| Spain | [Spanish Agency For Consumer Affairs, Food Safety and Nutrition](http://aesan.msssi.gob.es/en/AESAN/web/sobre_aesan/sobre_aecosan.shtml) |
| Switzerland | [Federal Food Safety and Veterinary Office](http://www.blv.admin.ch/index.html?lang=en) [Swiss Society for Nutrition](http://www.sge-ssn.ch/de/ich-und-du/essen-und-trinken/ausgewogen/lebensmittelpyramide/) |
| The former Yugoslav Republic of Macedonia | [Institute of Public Health](http://www.iph.mk/) |
| Turkey | [Ministry of Health](http://www.saglik.gov.tr/EN/ana-sayfa/2-0/20141211.html) |
| United Kingdom | [National Health Service](http://www.nhs.uk/Livewell/Goodfood/Pages/eatwell-plate.aspx) |
| Others | |
| Canada | [Health Canada](http://www.hc-sc.gc.ca/index-eng.php) |
| United States | [United States Department of Agriculture](http://www.usda.gov/wps/portal/usda/usdahome) |
| Chile | [Institute of Nutrition and Food Technology (INTA), the University](http://www.inta.cl/) [of Chile](http://www.inta.cl/) |
| Mexico | [National Institute of Public Health](http://www.insp.mx/) |
| Australia | [National Health and Medical Research Council](http://www.nhmrc.gov.au/your-health/nutrition) |

**Source:** http://www.fao.org/nutrition/education/food-based-dietary-guidelines/en/ [08.11.18]

Table A. 2 List of relevant databases, institutions and professional associations reporting on CPGs

| Relevant databases | (international level) |
| --- | --- |
| G-I-N | Guidelines International Network’s database |
| Guideline Central |  |
| NGC | US National Guideline Clearinghouse (until 09/18) |
| Relevant institutions | and expert associations (supranational level, selected countries) |
| WHO | World Health Organization |
| Europe |  |
| EAACI | European Academy of Allergy and Clinical Immunology |
| ESPEN | European Society for Clinical Nutrition and Metabolism |
| ESPGHAN | European Society for Pediatric Gastroenterology Hepatology and Nutrition |
| NICE | National Institute for Health and Care Excellence |
| The Americas |  |
| CDC | Centers for Disease Control and Prevention |
| ODPHP | Office of Disease Prevention and Health Promotion |
| USDA | United States Department of Agriculture. Center for Nutrition Policy and Promotion |
| ASPEN | American Society for Parenteral and Enteral Nutrition |
| AAAAI | The American Academy of Allergy, Asthma & immunology |
| AHS | Alberta Health Services |
| CSACI | Canadian Society of Allergy and Clinical Immunology |
| Australia |  |
| NWS | Nutrition Society of Australia |
| Relevant institutions | and expert association (national level) |
| ABAP | German Action Alliance for Allergy Prevention (Aktionsbündnis Allergieprävention) |
| AeDA | German Medical Association of Allergologists (Ärzteverband Deutscher Allergologen) |
| ADP | German Working Group on Dermatological Prevention (Arbeitsgemeinschaft Dermatologische Prävention) |
| AK-DIDA | German Task force on Dietetics in Allergology |
| AWMF | Arbeitsgemeinschaft der Wissenschaftlichen Medizinischen Fachgesellschaften |
| BVDD | Professional Association of German Dermatologists (Berufsverband der Deutschen Dermatologen) |
| BVHNO | German Professional Association of ENT Physicians (Berufsverband der HNO- Ärzte) |
| BVKJ | German Professional Association of Pediatricians (Berufsverband der Kinder- und Jugendärzte) |
| DAAB | German Allergy and Asthma Association (Deutscher Allergie- und Asthmabund) |
| DDG | German Dermatological Society (Deutsche Dermatologische Gesellschaft) |
| DGE | German Nutrition Society (Deutsche Gesellschaft für Ernährung) |
| DGEM | German Society for Nutritional Medicine (Deutsche Gesellschaft für Ernährungsmedizin) |
| DGHNOKHC | German Society for Oto-Rhino-Laryngology, Head and Neck Surgery (Deutsche Gesellschaft für Hals-Nasen-Ohren-Heilkunde, Kopf- und Hals-Chirurgie) |
| DGP | German Society for Pneumology (Deutsche Gesellschat für Pneumologie) |
| DGPM | German Society for Psychosomatic Medicine (Deutsche Gesellschaft für Psychosomatische Medizin) |
| GPA | German Society for Pediatric Allergology and Environmental Medicine (Gesellschaft für Pädiatrische Allergologie und Umweltmedizin) |
| GPGE | German Society for Pediatric Gastroenterology and Nutrition (Gesellschaft für pädiatrische Gastroenterologie und Ernährung) |

Table A. 3 Title, characteristics, and quality appraisal of included CPGs and FBDGs

| Citation | YoP | Lead | Full Title | Country/ | AGREE II | | | | | | |
| --- | --- | --- | --- | --- | --- | --- | --- | --- | --- | --- | --- |
|  |  | Association |  | Scope | (1) | (2) | (3) | (4) | (5) | (6) | Overall |
|  |  |  | CPGs & FBDGs |  | 66 % | 50 % | 42 % | 79 % | 24 % | 38 % | 63 % |
|  |  |  | CPGs only |  | 66 % | 55 % | 48 % | 82 % | 29 % | 50 % | 69 % |
|  |  |  | Allergy Prevention |  |  |  |  |  |  |  |  |
| [18] | 2013 | CPS | Dietary exposures and allergy prevention in high-risk infants: A joint statement with the Canadian Society of Allergy and Clinical Immunology | Canada | 72 % | 31 % | 26 % | 83 % | 17 % | 0 % | 42 % |
| [33] | 2014 | DGAKI & DGKJ^#^ | S3-Leitlinie Allergieprävention - Update 2014^#^ | Germany | 78 % | 58 % | 67 % | 81 % | 44 % | 96 % | 92 % |
| [20] | 2016 | Unclear | Guidelines for allergy prevention in Hong Kong | Hong Kong | 42 % | 8 % | 14 % | 83 % | 10 % | 46 % | 42 % |
| [21] | 2016 | SIPPS & SIAIP | Prevention of food and airway allergy: consensus of the Italian Society of Preventive and Social Paediatrics, the Italian Society of Paediatric Allergy and Immunology, and Italian Society of Pediatrics | Italy | 72 % | 44 % | 50 % | 92 % | 23 % | 42 % | 67 % |
| [34] | 2017 | Unclear | Dietary primary prevention of allergic diseases in children: the Philippine guidelines | Philippines | 64 % | 69 % | 51 % | 89 % | 15 % | 42 % | 67 % |
|  |  |  | Asthma |  |  |  |  |  |  |  |  |
| [51] | 2011 | GINA | Global strategy for the diagnosis and management of asthma in children 5 years and younger | International | 86 % | 31 % | 34 % | 75 % | 21 % | 4 % | 58 % |
| [27] | 2019 | SIGN & BTS | British Guideline on the management of asthma | United Kingdom | 92 % | 94 % | 81 % | 97 % | 38 % | 50 % | 92 % |
| [46] | 2020 | GINA | Global Strategy for Asthma Management and Prevention | International | 78 % | 56 % | 64 % | 94 % | 90 % | 75 % | 83 % |
|  |  |  | Atopic Eczema |  |  |  |  |  |  |  |  |
| [19] | 2011 | SIGN | Management of atopic eczema in primary care. | United  Kingdom | 83 % | 86 % | 73 % | 94 % | 40 % | 75 % | 92 % |
| [47] | 2013 | AAD | Guidelines of care for the management of atopic dermatitis: Section 1. Diagnosis and assessment of atopic dermatitis | United States | 86 % | 56 % | 64 % | 83 % | 17 % | 71 % | 83 % |
| [28] | 2018 | EDF | Consensus-based European guidelines for treatment of atopic eczema (atopic dermatitis) in adults and children: part I | European | 50 % | 86 % | 57 % | 92 % | 21 % | 46 % | 83 % |
| [37] | 2019 | AAP | The Effects of Early Nutritional Interventions on the Development of Atopic Disease in Infants and Children: The Role of Maternal Dietary Restriction, Breastfeeding, Hydrolyzed Formulas, and Timing of Introduction of Allergenic Complementary Foods | United States | 58 % | 42 % | 27 % | 56 % | 6 % | 71 % | 50 % |
| [41] | 2019 | CDA | Approach to the Assessment and Management of Pediatric Patients With Atopic Dermatitis: A Consensus Document. Section III: Treatment Options for Pediatric Atopic Dermatitis | Canada | 64 % | 36 % | 17 % | 72 % | 27 % | 88 % | 50 % |
| [52] | 2019 | Unclear | Guidelines on Management of Atopic Dermatitis in India: An Evidence-Based Review and an Expert Consensus | India | 69 % | 36% | 43 % | 83 % | 13 % | 58 % | 67 % |
|  |  |  | Food Allergies |  |  |  |  |  |  |  |  |
| [22] | 2010 | NIAID | Guidelines for the diagnosis and management of food allergy in the United States: report of the NIAID-sponsored expert panel | United States | 81 % | 89 % | 70 % | 83 % | 33 % | 67 % | 75 % |
| [26] | 2010 | BSACI | British Society for Allergy and Clinical Immunology guidelines for the management of egg allergy | United  Kingdom | 61 % | 44 % | 36 % | 69 % | 29 % | 25 % | 67 % |
| [29] | 2010 | AMS | Academy of medicine, Singapore-Ministry of Health clinical practice guidelines: management of food allergy | Singapore | 61 % | 81 % | 35 % | 81 % | 15 % | 0 % | 50 % |
| [48] | 2014 | EAACI | EAACI food allergy and anaphylaxis guidelines. Primary prevention of food allergy | European | 67 % | 78 % | 79 % | 78 % | 63 % | 88 % | 83 % |
| [40] | 2014 | AAAAI | Food allergy: a practice parameter update-2014 | United States | 28 % | 42 % | 53 % | 83 % | 25 % | 54 % | 75 % |
| [30] | 2017 | JSPACI | Japanese guidelines for food allergy 2017 | Japan | 31 % | 28 % | 13 % | 78 % | 21 % | 33 % | 58 % |
| [53] | 2017 | AAAAI | International consensus guidelines for the diagnosis and management of food protein-induced enterocolitis syndrome (FPIES): Executive summary-Workgroup Report of the Adverse Reactions to Foods Committee, American Academy of Allergy, Asthma & Immunology | International | 58 % | 56 % | 49 % | 86 % | 27 % | 54 % | 75 % |
| [38] | 2017 | BSACI | BSACI guideline for the diagnosis and management of peanut and tree nut allergy | United  Kingdom | 61 % | 39 % | 44 % | 72 % | 44 % | 42 % | 50 % |
| [35] | 2017 | NIAID | Addendum guidelines for the prevention of peanut allergy in the United States: Report of the National Institute of Allergy and Infectious Diseases-sponsored expert panel | United States | 78 % | 72 % | 52 % | 92 % | 31 % | 33 % | 83 % |
|  |  |  | FBDGs only |  | 67 % | 43 % | 31 % | 73 % | 15 % | 17 % | 52 % |
|  |  |  | Nutrition |  |  |  |  |  |  |  |  |
| [32] | 2010 | ÖGKJ | Österreichische Beikostempfehlungen | Austria | 64 % | 39 % | 33 % | 86 % | 27 % | 0 % | 58 % |
| [39] | 2012 | NHMRC | Infant Feeding Guidelines | Australia/ New Zealand | 72 % | 53 % | 55 % | 94 % | 8 % | 29 % | 58 % |
| [36] | 2013 | NHMRC | Australian Dietary Guidelines | Australia/ New Zealand | 97 % | 81 % | 54 % | 50 % | 35 % | 46 % | 58 % |
| [24] | 2014 | Health Canada, CPS, Dietitians of Canada and BCC | Nutrition for Healthy Term Infants: Recommendations from Six to 24 Months | Canada | 47 % | 28 % | 11 % | 81 % | 10 % | 0 % | 42 % |
| [54] | 2014 | Nordic Council of Ministers | Nordic Nutrition Recommendations 2012 | Scandinavia/ Iceland | 69 % | 31 % | 50 % | 61 % | 0 % | 25 % | 42 % |
| [17] | 2015 | AHS | Nutrition Guideline Healthy Infants and Young Children Introduction of Complementary Foods | Canada | 78 % | 28 % | 19 % | 78 % | 8 % | 4 % | 33 % |
| [25] | 2015 | Health Canada, CPS, Dietitians of Canada and BCC | Nutrition for healthy term infants: Recommendations from birth to six months | Canada | 47 % | 28 % | 11 % | 67 % | 8 % | 0 % | 33 % |
| [15] | 2016 | NGIL (part of the BZfE) | Ernährung und Bewegung von Säuglingen und stillenden Frauen | Germany | 81 % | 50 % | 33 % | 67 % | 4 % | 38 % | 50 % |
| [23] | 2017 | Unclear | Feeding Guidelines for Infants and Young Toddlers: A Responsive Parenting Approach | United States | 69 % | 56 % | 35 % | 78 % | 27 % | 4 % | 58 % |
| [55] | 2018 | NGIL  (part of the BZfE) | Diet and Lifestyle Before and During Pregnancy - Practical Recommendations of the Germany-wide Healthy Start - Young Family Network | Germany | 72 % | 56 % | 41 % | 78 % | 2 % | 42 % | 67 % |
| [31] | 2019 | THL | EATING TOGETHER - food recommendations for families with children | Scandinavia/ Iceland | 67 % | 53 % | 23 % | 72 % | 23 % | 29 % | 67 % |
| [16] | 2019 | SGE | Ernährung des Säuglings im ersten Lebensjahr | Switzerland | 44 % | 8 % | 10 % | 50 % | 13 % | 0 % | 42 % |
|  |  |  | Food Allergies |  |  |  |  |  |  |  |  |
| [56] | 2019 | Unclear | Ernährungsberatung bei Kindern mit IgE-vermittelten Nahrungsmittelallergien | Switzerland | 61 % | 47 % | 29 % | 89 % | 31 % | 4 % | 67 % |

YoP: Year of publication

CPS, Canadian Paediatric Society; DGAKI, Deutsche Gesellschaft für Allergologie und klinische Immunologie; DGKJ, Deutsche Gesellschaft für Kinder- und Jugendmedizin; SIPPS, Italian Society of Preventive and Social Paediatrics; SIAIP, Italian Society of Paediatric Allergy and Immunology; GINA, Global Initiative for Asthma; SIGN, Scottish Intercollegiate Guidelines Network; BTS, British Thoracic Society; AAD, American Academy of Dermatology; EDF, European Dermatology Forum; AAP, American Academy of Pediatrics; CDA, Canadian Dermatology Association; NIAID, National Institute of Allergy and Infectious Diseases; BSACI, British Society for Allergy and Clinical Immunology; AMS, Academy of medicine, Singapore; EAACI, European Academy of Allergy and Clinical Immunology; AAAAI, American Academy of Allergy, Asthma and Immunology; JSPACI, Japanese Society of Pediatric Allergy and Clinical Immunology; ÖGKJ, Österreichische Gesellschaft für Kinder- und Jugendheilkunde; NHMRC, National Health and Medical Research Counsil; AHS, Alberta Health Services; BCC, Breastfeeding Committee for Canada; NGIL, Netzwerk Gesund ins Leben – eine Initiative von IN FORM; BZfE, Bundeszentrum für Ernährung; THL, Finnish Institute for Health and Welfare, Finland; SGE, Schweizerische Gesellschaft für Ernährung

\# This German S3-guideline allergy prevention was not valid at the time of search, but was included in the sample because it is the most comprehensive German guideline on allergy prevention

Table A. 4 Recommendation statements on the introduction of complementary foods and allergenic foods in high-risk infants

| Recommendation statement | Recommended | | Not recommended | | No recommendation made | | SoR | LoE |
| --- | --- | --- | --- | --- | --- | --- | --- | --- |
|  | CPG | FBDG | CPG | FBDG | CPG | FBDG |  |  |
| Introducing solid foods at  4-6 months of age |  | [15] |  |  |  |  |  |  |
| Introducing common food | [28]^‡^ |  |  |  |  |  | ++ | ++ |
| allergens from 6 months of age |  | [24]^‡^ |  |  |  |  |  |  |
|  |  | [25]^‡^ |  |  |  |  |  |  |
| Delayed introduction of potential food allergens |  |  |  | [25]^‡^ |  |  |  |  |
| Introducing peanut at 4-6  months of age | [35] |  |  |  |  |  | +++ | ++ |
| Introducing peanut from 6 | [35] |  |  |  |  |  | + | + |
| months of age | [37] |  |  |  |  |  |  |  |
| Early introduction of peanut |  |  |  |  | [38] |  |  |  |
|  |  |  |  |  | [46] |  |  |  |
| Consult health care providers | [35] |  |  |  |  |  |  |  |
| and approach each case on an |  | [24] |  |  |  |  |  |  |
| individual basis |  | [23] |  |  |  |  |  |  |
| Seek guidance during weaning, to  ensure nutritional adequacy | [53] |  |  |  |  |  | +++ | ++ |

SoR, Strength or recommendation; LoE, Level of evidence

^‡^ recommendation applies to children at average and at high risk for allergies
